## Extended Methods and Figures for "Identifying proteomic risk factors for cancer using prospective and exome analyses: 1,463 circulating proteins and risk of 19 cancers in the UK Biobank"

### Extended Methods: cancer specific adjustments

All cancer-specific multivariable regression models were stratified by age and sex and adjusted for region and Townsend deprivation index (minimally adjusted model) and additionally adjusted for the following for:

***Bladder cancer*** - [smoking status and number of cigarettes smoked (8 categories: never, former smoker and < 15 cigarettes/day, former smoker and ≥15 cigarettes/day, former smoker and number of cigarettes smoked unknown, current smoker and < 15 cigarettes/day, current smoker and ≥15 cigarettes/day, current smoker and number of cigarettes smoked unknown, unknown), cigarette-pack years (5 categories: quintiles, unknown), and BMI (4 categories <25, 25-29, 30-34, ≥35 kg/m^2^)].

***Breast cancer -*** [family history of breast cancer (yes, no or unknown), parity and age at first birth (11 categories: ≥3 kids, <25 years, ≥3 kids, 25-29 years, ≥3 kids, ≥30 years, ≥3 kids, age not reported, : 1-2 kids, <25 years, 1-2 kids, 25-29 years, 1-2 kids, ≥30 years, 1-2 kids, age not reported, no children, not applicable, men), age menarche (6 categories: <11 years, 12-13 years, 14-15 years, ≥16 years, men, unknown), hormone replacement therapy use (4 categories: never, past, men, unknown), oral contraceptive use (5 categories: never, for <20 years, for ≥20 years, unknown, men), alcohol intake (6 categories: < 1 g/day, 1-9 g/day, 10-19 g/day, ≥20 g/day, non-drinkers, unknown), physical activity (6 categories: <10 metabolic equivalent (MET) hours per week, 10-19 MET hours per week, 20-39 MET hours per week, 40-59 MET hours per week, ≥60 MET hours per week, unknown), BMI (4 categories), menopausal status (4 categories: pre-menopausal, post-menopausal, men, unknown), and an interaction between BMI (4 categories) and menopausal status (4 categories)].

***Head and neck cancer (including subtypes) and liver cancer -*** [smoking status (6 categories: never, former smoker, current smoker, <15 cigarettes/day, current smoker, ≥15 cigarettes/day, current smoker, number of cigarettes unknown, unknown), BMI (4 categories), and alcohol intake (6 categories)];kidney cancer [smoking status and number of cigarettes smoked (8 categories), cigarette-pack years (5 categories), and BMI (4 categories)]; leukemia [smoking status and number of cigarettes smoked (8 categories)];

***Lung cancer (including subtypes) -*** [family history of lung cancer (yes, no or unknown), smoking status and number of cigarettes smoked (8 categories), smoking status and years of smoking (9 categories: never, former smoker and < 30 years, former smoker and ≥30 years, former smoker and number of cigarettes smoked unknown, current smoker and < 30 years, current smoker and ≥ 30 years, current smoker and number of cigarettes smoked unknown, unknown status, unknown), and particulate matter (5 categories: quintiles, unknown)].

***Melanoma -*** [use sun or UV protection (6 categories: never/rarely, sometimes, most of the time, always, do not go out in the sunshine, unknown), hours spent outdoors on a summer day (continuous), ease of skin tanning (5 categories: never tan only burn, get mildly or occasionally tanned, get moderately tanned, get very tanned, unknown)]; for colorectal cancer and its subtypes [family history of colorectal cancer (yes, no or unknown), waist to hip ratio (6 categories: quintiles, unknown), frequency of processed meat intake (< once per week, ≥ once per week, unknown), physical activity (6 categories), BMI (4 categories), alcohol intake (6 categories), and smoking status and years of smoking (9 categories)].

***Ovarian cancer -*** [family history of breast cancer (yes, no or unknown), parity and age at first birth (11 categories), age menarche (6 categories), hormone replacement therapy use (4 categories), oral contraceptive use (5 categories, smoking status and years of smoking (9 categories), BMI (4 categories), menopausal status (4 categories), and an interaction term between BMI (4 categories) and menopausal status (4 categories)].

pancreas cancer - [smoking status and number of cigarettes smoked (8 categories), and BMI (4 categories)];

prostate cancer - [family history of prostate cancer (yes, no or unknown), and body mass index (BMI) (4 categories).

***Stomach cancer -*** [BMI (4 categories), and alcohol intake (6 categories)].

***Thyroid cancer -*** [BMI (4 categories)].

***Uterine cancer*** [parity and age at first birth (11 categories), age menarche (6 categories), hormone replacement therapy use (4 categories), oral contraceptive use (5 categories, physical activity (6 categories), smoking status and years of smoking (9 categories), BMI (4 categories), and menopausal status (4 categories)].

We did not adjust brain cancer and the blood cancer sub-types other than leukemia analyses beyond the minimally adjusted model since there were no known relevant confounders in this dataset.

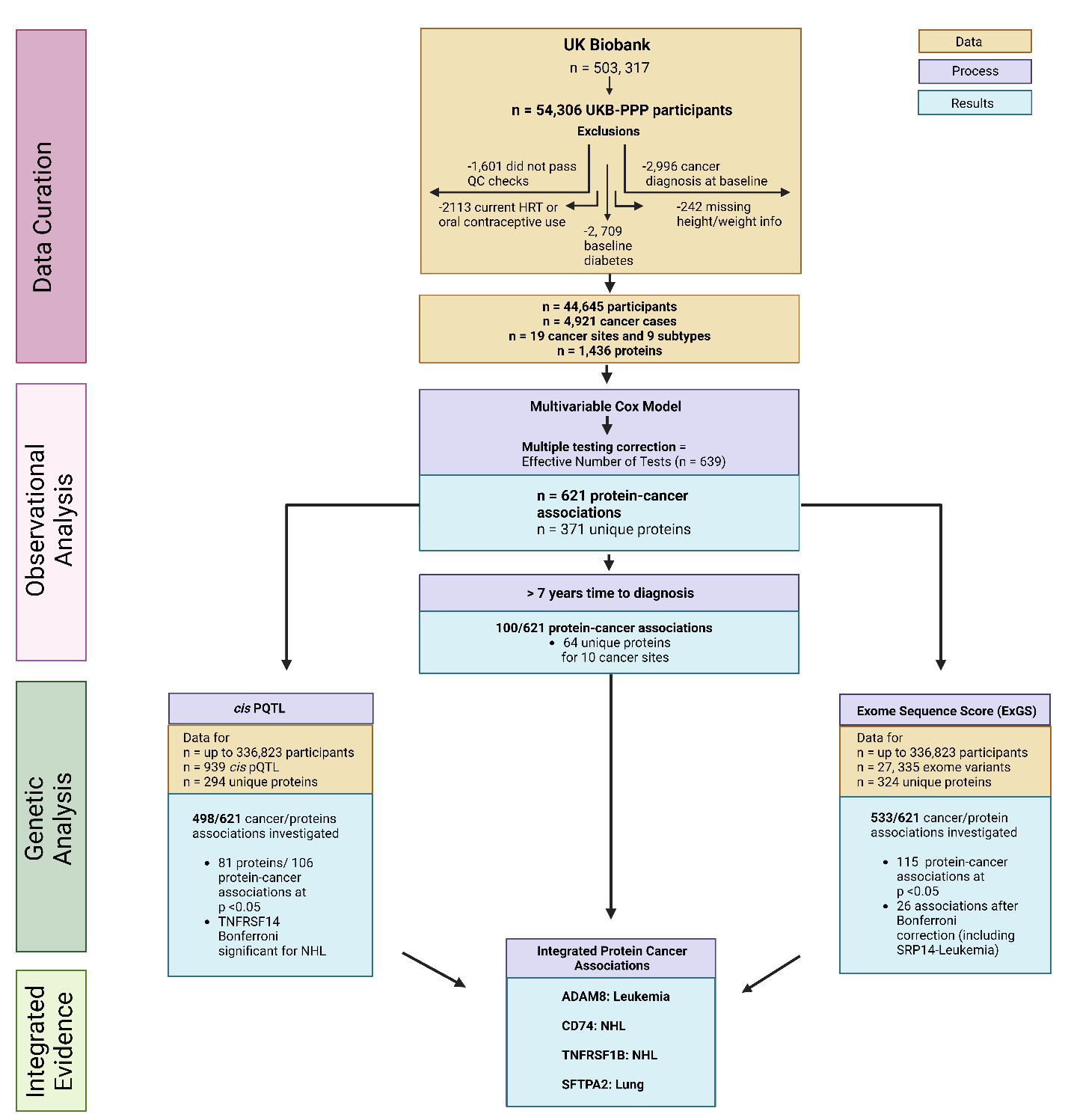

### Extended Figure 1. Study design flow chart and results summary

Multivariable Cox Model was performed on individuals that were part of the UK Biobank Pharma Proteomics Project (UKB-PPP) after excluding individuals that failed QC, had missing data, were diabetic or were using hormone replacement therapy (HRT) or using oral conceptive use at baseline. Genetic evidence using cis-pQTLs and exome sequencing scoring (ExGS) for protein-cancer associations was investigated. Lastly, data was integrated to investigate protein-cancer associations with multiple levels of support.

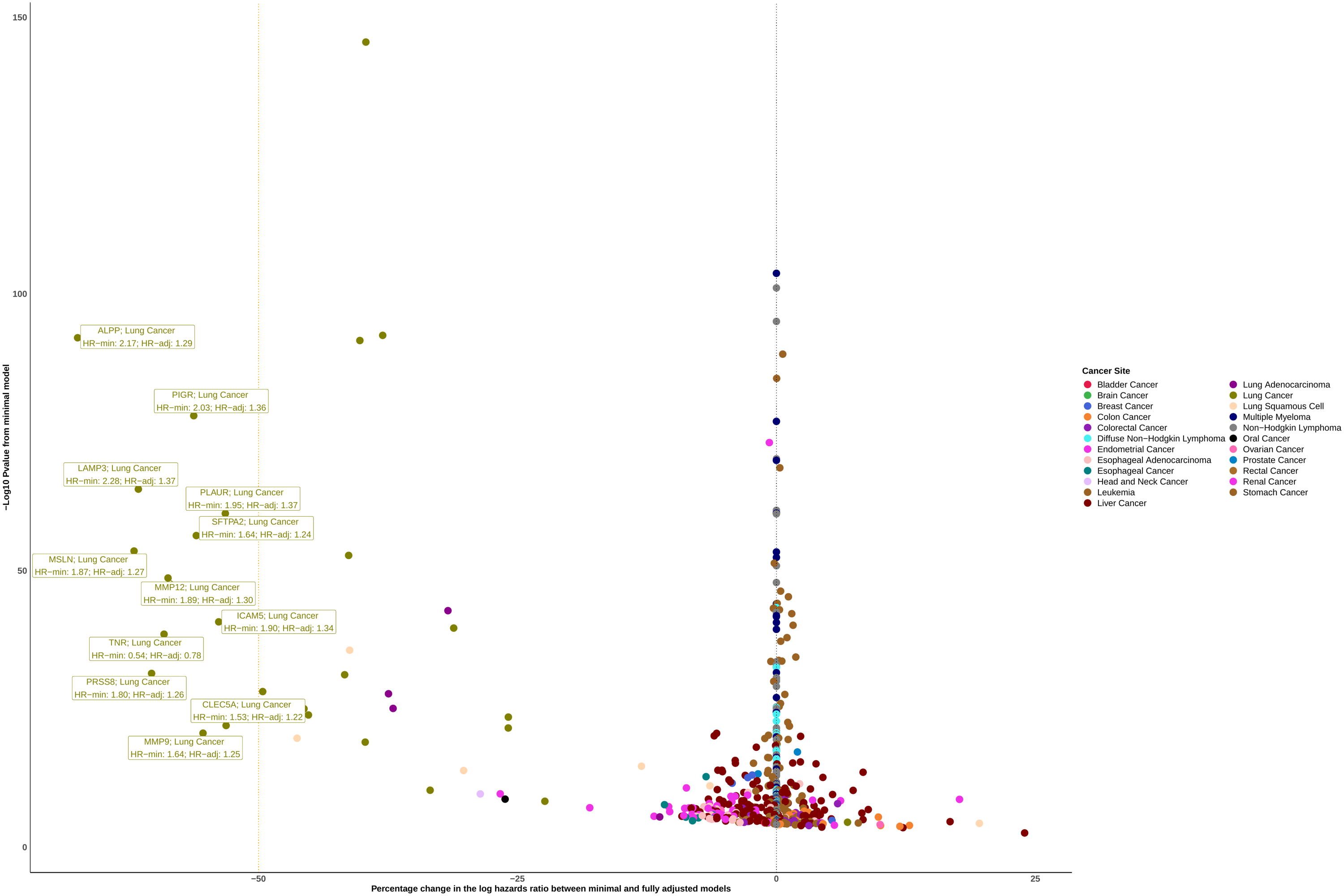

### Extended Figure 2. Percentage change in the log hazard ratios between fully and minimally adjusted models

This figure displays the differences between the minimally model and fully adjusted models with the x-axis displaying the percentage change with the y-axis displaying the -log10(P-value) from the minimal model. The colours represent the cancer site.
